# Poor Surgical Outcomes Following *Paenibacillus* Infant Infectious Hydrocephalus

**DOI:** 10.1101/2025.05.08.25327256

**Authors:** Jessica E. Ericson, Davis Natukwatsa, Peter Ssenyonga, Justin Onen, John Mugamba, Ronald Mulondo, Sarah U. Morton, Mercedeh Movassagh, Kelsey Templeton, Christine Hehnly, Edith Mbabazi-Kabachelor, Abhaya V. Kulkarni, Benjamin C. Warf, James R. Broach, Joseph N. Paulson, Steven J. Schiff

## Abstract

**Objective:** We previously identified *Paenibacillus species* in the cerebrospinal fluid of 44% of infants presenting for neurosurgical evaluation with findings consistent with postinfectious hydrocephalus (PIH) in Eastern Uganda. Here we sought to compare outcomes among hydrocephalic infants with and without *Paenibacillus* detection at the time of hydrocephalus surgery.

**Methods:** In a prospective observational study, 78 infants with PIH who underwent a cerebrospinal fluid (CSF) diversion prior to 90 days of age had a positive CSF polymerase chain reaction result for *Paenibacillus* species (PP), and 111 had a negative result (PN). The primary outcome was diversion failure-free survival defined as being alive without diversion failure at last patient contact. Secondary outcomes included overall survival and diversion success.

**Results:** After a median follow-up period of 35.7 months, the primary outcome was observed in 42 PP patients (54%) and in 76 PN patients (68%) (adjusted hazard ratio (aHR), 2.45; 95% confidence interval [CI], 1.42 to 4.22; P=0.001). PP patients who underwent endoscopic diversion had the worst primary event rate (aHR, 6.47; 95% CI, 2.40 to 17.42; P<0.001). Death from any cause occurred in 16 PP patients (20%) and 9 PN patients (8%) (aHR, 3.47; 95% CI, 1.44 to 8.37; P=0.006). Diversion failure occurred in 28 PP patients (36%) and in 29 PN patients (26%) (aHR, 2.24; 95% CI, 1.31 to 3.85; P=0.003).

**Conclusions:** In this study, *Paenibacillus* detection in the CSF at the time of hydrocephalus surgery was associated with a significantly increased rate of the composite of diversion failure or death, death, and diversion failure, and was particularly increased for patients who had an endoscopic diversion.

## Introduction

Post-infectious hydrocephalus (PIH) is the most common cause of hydrocephalus in low-resource settings.^1^ PIH most often occurs as a complication of neonatal sepsis and meningitis. Most cases of PIH require neurosurgery to arrest the abnormal accumulation of. Because of the primary insult to the brain from the original infection in children with PIH, such disabilities are common even after successful treatment of the hydrocephalus.^2^

Since the mid-20th century, the standard of care for hydrocephalus management has been placement of ventriculoperitoneal shunts (VPS).^3^ Children with a VPS require ongoing specialized neurosurgical care and frequently require additional surgeries to manage shunt failure or infection. Endoscopic third ventriculostomy (ETV) with or without choroid plexus cauterization (CPC) has been introduced as an alternative means of managing hydrocephalus that does not have the risks associated with implanted hardware^3^. Randomized clinical trials have demonstrated that developmental outcomes, treatment failure and brain growth after ETV/CPC are similar to VPS.^4–6^ As a consequence, ETV/CPC is often the preferred neurosurgical procedure for management of infant PIH, especially in low-resource settings. Several scoring systems have been developed to predict which patients are likely to have their hydrocephalus successfully managed with an ETV.^7^ The score that performs the best in infants cared for in low-resource settings is the Uganda ETV success score.^7^ However, even patients who are predicted to have success with an ETV sometimes experience failure and ultimately require VPS placement for management of their hydrocephalus. When ETV/CPC fails, reopening of a closed ETV can sometimes provide long-term shunt freedom, but many cases of ETV/CPC failure ultimately require shunt placement.^8^

*Paenibacillus species* have previously been associated with sporadic infections in adults but have more recently been described as a cause of meningitis and postinfectious hydrocephalus in infants.^9,10^ These organisms are spore-forming Gram-positive bacteria that have been found throughout the environment in soil, freshwater lakes, honeybee colonies and in the bowel flora of healthy adults.^11,12^ We recently identified *Paenibacillus species* in the cerebrospinal fluid of 44% of infants presenting for neurosurgical evaluation in Eastern Uganda who had findings consistent with PIH.^13^ The initial description included 209 patients with PIH and focused on the molecular methods used to identify this novel putative pathogen. This report provides a clinical comparison of infant neurosurgical outcomes following diversion surgery in *Paenibacillus*-associated PIH and PIH due to other causes.

## Methods

### Study Oversight

This study was a single center observational prospective study conducted at CURE Children’s Hospital of Uganda. The study design and a description of the overall cohort has previously been published.^13,14^ The study protocol was approved by the Institutional Review Boards at CURE Children’s Hospital of Uganda with oversight of the Ugandan National Council on Science and Technology, The Pennsylvania State University, and Yale University. This analysis was conducted following STROBE guidelines for observational studies.

### Study Population

The original study included 400 infants younger than 90 days old with hydrocephalus treated at CURE Children’s Hospital of Uganda in 2016 and 2017 (Figure 1).^13,14^ Infants were considered to have non-postinfectious hydrocephalus (NPIH) if they had a congenital malformation noted on imaging (i.e.: Dandy-Walker cyst) or were born with macrocephaly, and were considered to have PIH if the infant was born with a normal head size and had a febrile episode or seizures prior to the onset of macrocephaly. All patients had cerebrospinal fluid (CSF) collected for molecular analysis. Metagenomic sequencing revealed *Paenibacillus thiaminolyticus* as a common pathogen leading us to perform quantitative PCR tests for both *P. thiaminolyticus* and *Paenibacillus* sp. on all patients who had sufficient CSF remaining to perform these tests.^13^ For this clinical analysis, we excluded patients with NPIH (N=191). Additionally, because this clinical analysis assesses the impact of *Paenibacillus* detection on diversion success, patients who did not undergo a CSF diversion procedure were excluded (N=20).

**Figure 1.**
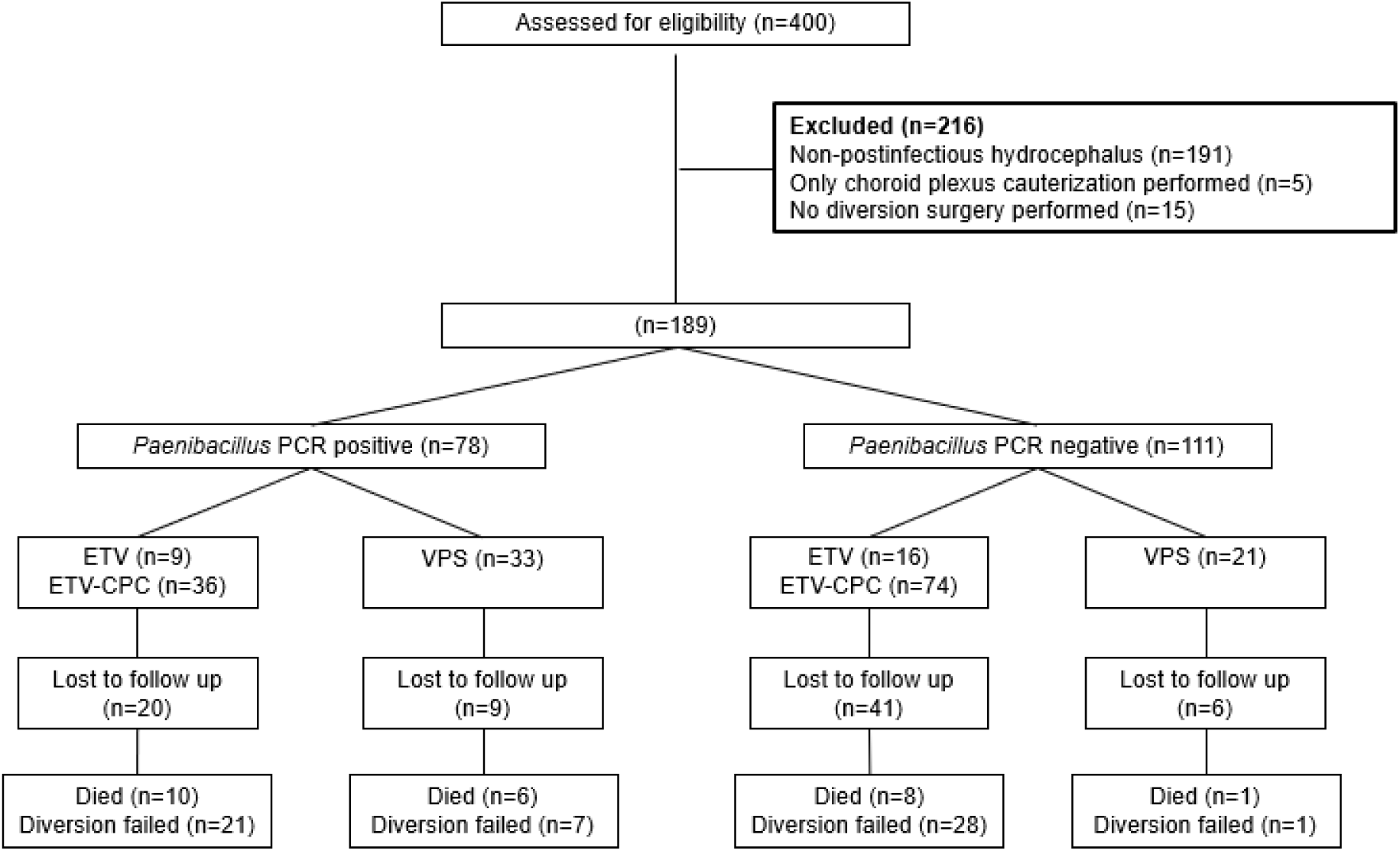
Patient flow diagram.

Patients were followed for routine care and additional neurosurgical procedures, if any, were recorded. The timing and type of follow-up evaluations, surgical procedures and medical therapies were determined by the treating clinician. For patients who underwent a head CT scan for surgical planning, each image was reviewed by two blinded investigators and the presence of loculations, abscess, calcifications and ventricular debris were noted. For those who died, the date of death and cause of death were recorded. We attempted to contact all participants by phone during the month of June 2023 to determine the vital status, dead vs. alive, and, for those who died, the date and cause of death.

### Definitions

Based on our prior work, patients with >33 reads/mL of *P. thiaminolyticus* or >55 reads/mL of *Paenibacillus* genus were considered to be *Paenibacillus* positive (PP); all other patients were considered to be *Paenibacillus* negative (PN).^13^ The primary diversion procedure was defined as the first occurrence of: VPS, ETV, or ETV/CPC. The primary diversion was considered to have failed when a primary diversion procedure was followed by: a second diversion procedure, ventricular washout, VPS revision or VPS removal. For patients who had an ETV or ETV/CPC as their first diversion surgery, we abstracted the basic and extended Uganda ETV success scores from the medical record. The basic method has previously been described with points assigned for each of: the cause of hydrocephalus, the patient’s age at the time of surgery and the extent of CPC performed.^7^ The second extended method adds additional points if the Aqueduct of Sylvius is closed or if the prepontine cistern is open (unscarred).^15,16^ Scores of 7-10 were considered to have a high probability of success, scores of 4-6 a moderate probability of success and scores of ≤3 a low probability of success.^7^

### Outcomes

The primary outcome was diversion failure-free-survival (FFS) defined as the time from the primary surgical diversion to diversion failure or death from any cause. Overall survival (OS) was defined as the time from surgical diversion to death from any cause. Diversion success was defined as being alive without additional neurosurgical procedures at the last known contact with the patient.

### Statistical Analysis

Patient characteristics, the type of surgery performed and head computed tomography (CT) results for PP and PN patients were compared using Fisher’s exact test for categorical variables and Wilcoxon rank sum test for continuous variables. We calculated the sensitivity, specificity and positive and negative predictive values of having at least 3 CT scan findings for predicting the *Paenibacillus* CSF PCR result.

Outcomes for infants with and without *Paenibacillus* detected were assessed utilizing various methodologies. The FFS, OS and diversion success rates were estimated using Kaplan Meier methodology for diversion type, along with 95% Confidence Intervals (CIs) calculated using the standard error derived from the Greenwood formula.^17^ Statistical hypothesis testing was conducted using the log-rank test to assess survival differences among the groups. No correction for multiple testing was applied. To discern differential rates between the two therapeutic cohorts, the 95% CI for the rate difference was computed using the normal approximation technique. Additionally, we used Cox proportional hazards regression models to compare outcomes for PP and PN infants, adjusted for age at the time of the diversion procedure and the type of diversion procedure (VPS vs. endoscopic diversion). Due to an insufficient number of events, we opted not to incorporate a term representing the interaction between diversion type and the *Paenibacillus* PCR result in our model. We used the Cochran-Mantel-Haenszel test to compare each outcome for patients with and without *Paenibacillus* detected after 6 and 12 months from the diversion procedure. Finally, we calculated “*Paenibacillus*-attributable harm” by determining the differences in the number of deaths after surgical diversion and the 1-year diversion failure among those who survived the first year between PN and PP normalized by the proportion of patients receiving VPS and ETV/CPC in each group.

As a sensitivity analysis, we subset our cohort to those patients with an endoscopic diversion as their primary diversion procedure and compared outcomes in patients with and without *Paenibacillus* detected. We analyzed this cohort similar to above, except that the Cox regression analysis was adjusted for the Uganda ETV success score tertile. Age was not included as a covariate in addition to the ETV success score due to the limited number of events and its inclusion in the Uganda ETV success score. A separate regression was performed for both the basic and extended Uganda ETV success score scales.

Finally, to detect the presence of active *Paenibacillus* infection in the hydrocephalic cohort, we performed RNA sequencing on CSF and blood samples. Organism alignment was conducted using Chan Zuckerberg ID (CZID)^18^ followed by the PathSeeker^19^ pathogen identification pipeline to identify PP samples. In brief, the decontam and topic modeling algorithms from the *PathSeeker* pipeline were both applied to differentiate between contaminants and true organisms. For a final soft filtering step, organisms lacking reads which aligned to proteins or with fewer than 100 nucleotide reads were removed from the dataset. A two-sided alpha level of 0.05 was employed as the threshold for determining statistical significance. Analyses were performed using R software (R Project for Statistical Computing) and Stata 14.2 (Stata Corp., College Station, Texas).

## Results

### Characteristics of PP and PN Patients

All of the 189 infants with PIH who underwent a diversion procedure from January 2016 to October 2019 had PCR for *Paenibacillus* performed on the CSF; 78 (41%) had a positive result. Three infants, 3/78 (4%) with a positive PCR also had a culture that grew *Paenibacillus*; all three of these were *P. thiaminolyticus*. PP infants had a younger median age and smaller median weight at the time of admission than PN patients, 58 days (25^th^, 75^th^ percentiles: 48, 69) vs. 68 days (56, 80), P<0.001, and 4600 grams (3900, 5400) vs. 5200 (4200, 6010), P=0.003, respectively (Table 1). PP infants were more likely to undergo one or more ventricular washout procedures prior to their first diversion surgery, 46/78 (59%) vs. 14/111 (13%), P<0.001. This resulted in PP patients being older than PN patients at the time of the diversion surgery, 94 days (69, 131) vs. 70 days (61, 84), P<0.001. PP patients were more likely to have been born at home rather than in a healthcare setting compared to PN patients, 41/78 (52%) vs. 39/111 (35%), respectively, P=0.025. Other patient characteristics were not different between the two groups and a similar proportion of infants in both groups had fever or seizures at admission. ETV/CPC was the most common diversion surgery performed, 109/189 (58%), followed by VPS, 54/189 (29%), and ETV, 26/189 (14%).

**Table 1.** Clinical characteristics of infants undergoing surgical management of post-infectious hydrocephalus due to *Paenibacillus species* vs. other causes.

|  | *Paenibacillus*<br>PCR Negative<br>N=111 (59%) | *Paenibacillus*<br>PCR Positive<br>N=78 (41%) | P |
| --- | --- | --- | --- |
| Median age at admission, days (25<sup>th</sup>, 75<sup>th</sup> percentiles) | 68 (56, 80) | 58 (48, 69) | <0.001 |
| ≤28 days | 1 (1) | 1 (1) |  |
| 29-59 days | 29 (26) | 39 (50) |  |
| 60-89 days | 70 (63) | 35 (45) |  |
| ≥90 days | 11 (10) | 3 (4) |  |
| Gestational age category |  |  | 0.442 |
| Preterm | 3 (3) | 4 (5) |  |
| Term | 105 (97) | 69 (94) |  |
| Sex |  |  | 1.000 |
| Female | 48 (43) | 34 (44) |  |
| Male | 63 (57) | 44 (56) |  |
| Delivery location |  |  | 0.025 |
| Healthcare setting | 72 (65) | 37 (47) |  |
| Home | 39 (35) | 41 (52) |  |
| Delivery mode |  |  | 1.000 |
| Vaginal | 107 (96) | 76 (97) |  |
| C-section | 4 (4) | 2 (3) |  |
| Median weight at presentation, grams<br>(25<sup>th</sup>, 75<sup>th</sup> percentiles) | 5.2 (4.2, 6.0) | 4.6, (3.9, 5.4) | 0.003 |
| Malaria smear result |  |  | 1.000 |
| Negative | 109 (99) | 78 (100) |  |
| Positive | 1 (1) | 0 (0) |  |
| HIV antibody test result |  |  | 0.334 |
| Negative | 100 (90) | 75 (96) |  |
| Positive | 4 (4) | 1 (1) |  |
| Unknown | 7 (6) | 2 (3) |  |
| Received antibiotics prior to admission |  |  | 0.059 |
| No | 73 (66) | 39 (50) |  |
| Yes | 27 (24) | 31 (40) |  |
| Unknown | 10 (9) | 8 (10) |  |
| Fever at presentation | 7 (6) | 4 (5) | 1.000 |
| Seizures at presentation | 15 (14) | 10 (13) | 1.000 |
| Head imaging findings |  |  |  |
| None | 23 (21) | 4 (5) | 0.003 |
| Abscess | 26 (24) | 37 (48) | 0.001 |
| Calcification | 50 (46) | 56 (73) | <0.001 |
| Loculations | 64 (59) | 66 (86) | <0.001 |
| Debris | 42 (39) | 46 (60) | 0.007 |
| All 4 CT findings | 15 (14) | 26 (34) | <0.001 |
| Median age at diversion surgery, days<br>(25<sup>th</sup>, 75<sup>th</sup> percentiles) | 70 (61, 84) | 94 (69, 131) | <0.001 |
| Median number of irrigations before<br>hydrocephalus surgery | 0 (0, 0) | 1 (0, 1) | <0.001 |
| Median number of surgeries, (25, 75<br>percentiles) | 1 (1, 2) | 2 (2, 3) | <0.001 |
| Type of diversion surgery performed |  |  | 0.002 |
| Ventriculoperitoneal shunt | 21(19) | 33 (42) |  |
| Endoscopic third ventriculostomy | 17 (15) | 9 (12) |  |
| ETV with choroid plexus cauterization | 73 (66) | 36 (46) |  |

Most patients had a head CT performed prior to neurosurgery, 185/189 (98%). PP infants were more likely than PN infants to have head CT findings indicative of sequelae of intracranial infection and a third of patients, 26/77 (34%), with *Paenibacillus* had all four features of abscess, loculations, calcifications and ventricular debris noted on head CT compared with just 15/108 (14%) of patients without *Paenibacillus* infection, P<0.001. In this cohort, the presence of >=3 abnormalities on head CT was 58% (51, 65) sensitive and 73% (67, 79) specific for *Paenibacillus* detection in the CSF with a positive predictive value of 60% (53, 67) and a negative predictive value of 71% (65, 77) (Supplemental Table 1).

### Association of Patient Characteristics and Outcomes

In separate unadjusted Cox regression analyses for each characteristic, age, sex, the presence of an abscess, calcifications, debris or loculations on head CT imaging, and having a ventricular washout procedure performed prior to diversion were not associated with FFS, OS and diversion success (Figure 2). The Uganda ETV success score and diversion surgery type were associated with FFS and diversion success but not OS; *Paenibacillus* PCR was associated with FFS and OS but not with diversion success.

**Figure 2.**
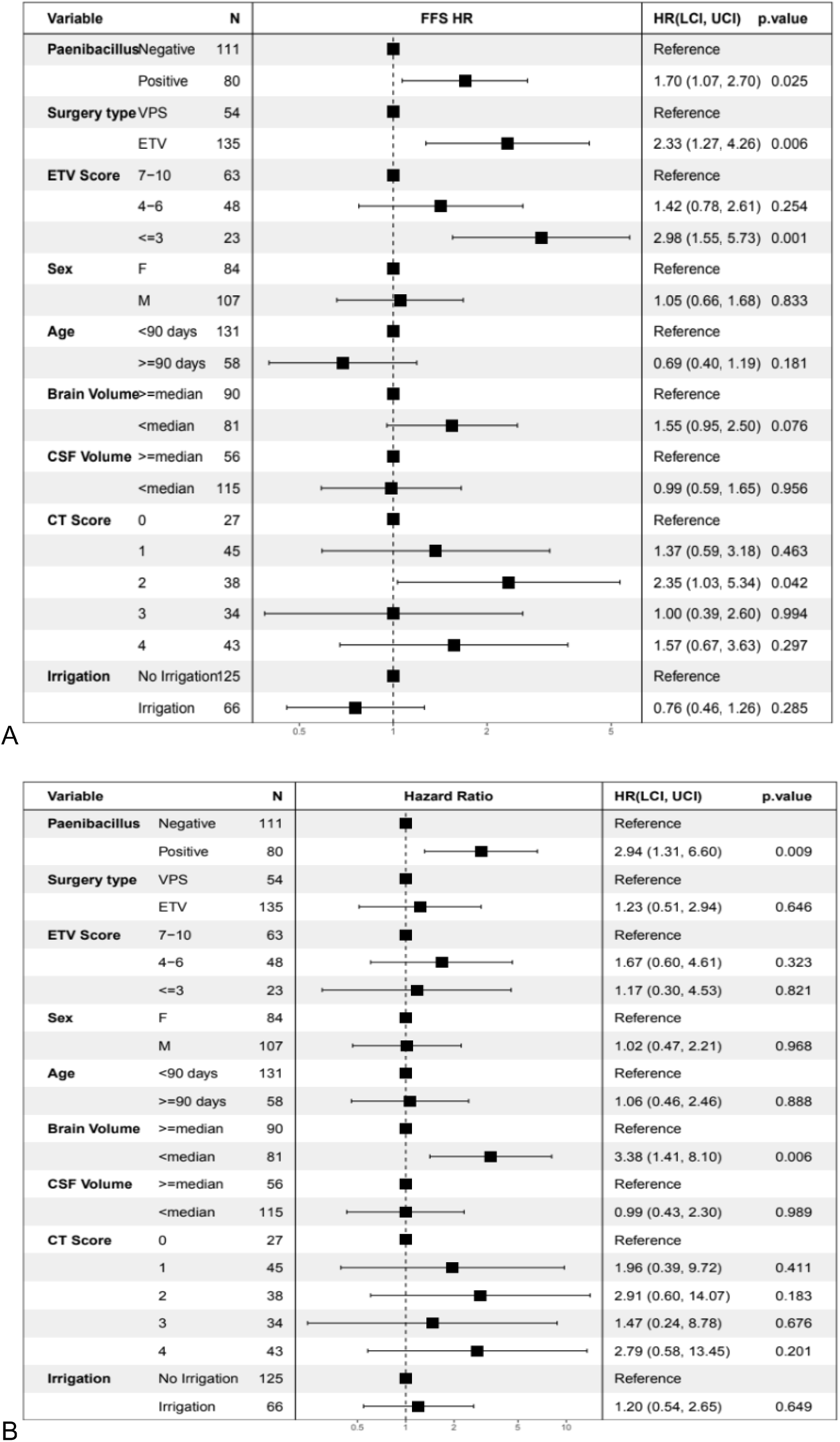
Univariate rate of diversion failure or death from any cause (A) and death from any cause (B) in subgroups.

### Outcomes

After a median follow-up period of 35.7 months, the primary outcome had occurred in 36 PP patients (46%) and in 35 PN patients (32%). FFS was most common among patients who underwent VPS (Supplemental Figure 1). When adjusted for age at the time of surgery and surgery type, *Paenibacillus* infection was associated with worse FFS, adjusted hazard ratio (aHR) =2.45 (1.42, 4.22) (Figure 3). This increase was predominantly experienced by patients who had *Paenibacillus* detected and had an endoscopic diversion performed as their primary diversion procedure, aHR=6.47 (2.40, 17.42). PP patients died more often than PN patients, 16/78 (20%) vs 9/111 (8%) following both types of surgical diversion and, when adjusted for age and surgery type, PP patients had worse OS than PN patients, aHR=3.47 (1.44, 8.37). PP patients who underwent endoscopic diversion had the worst survival, aHR=6.01 (1.36, 26.6) (Figure 4). Surgical CSF diversion was more likely to fail in PP patients compared to PN patients, aHR=2.24 (1.31, 3.85) and was particularly marked for patients who underwent an endoscopic diversion, aHR= 6.03 (2.16, 16.8) (Figure 5). Additionally, *Paenibacillus* positivity was associated with worse FFS at 12 months and diversion success at 6 and 12 months when ETV or ETV/CPC was performed (Table 2).

**Figure 3.**
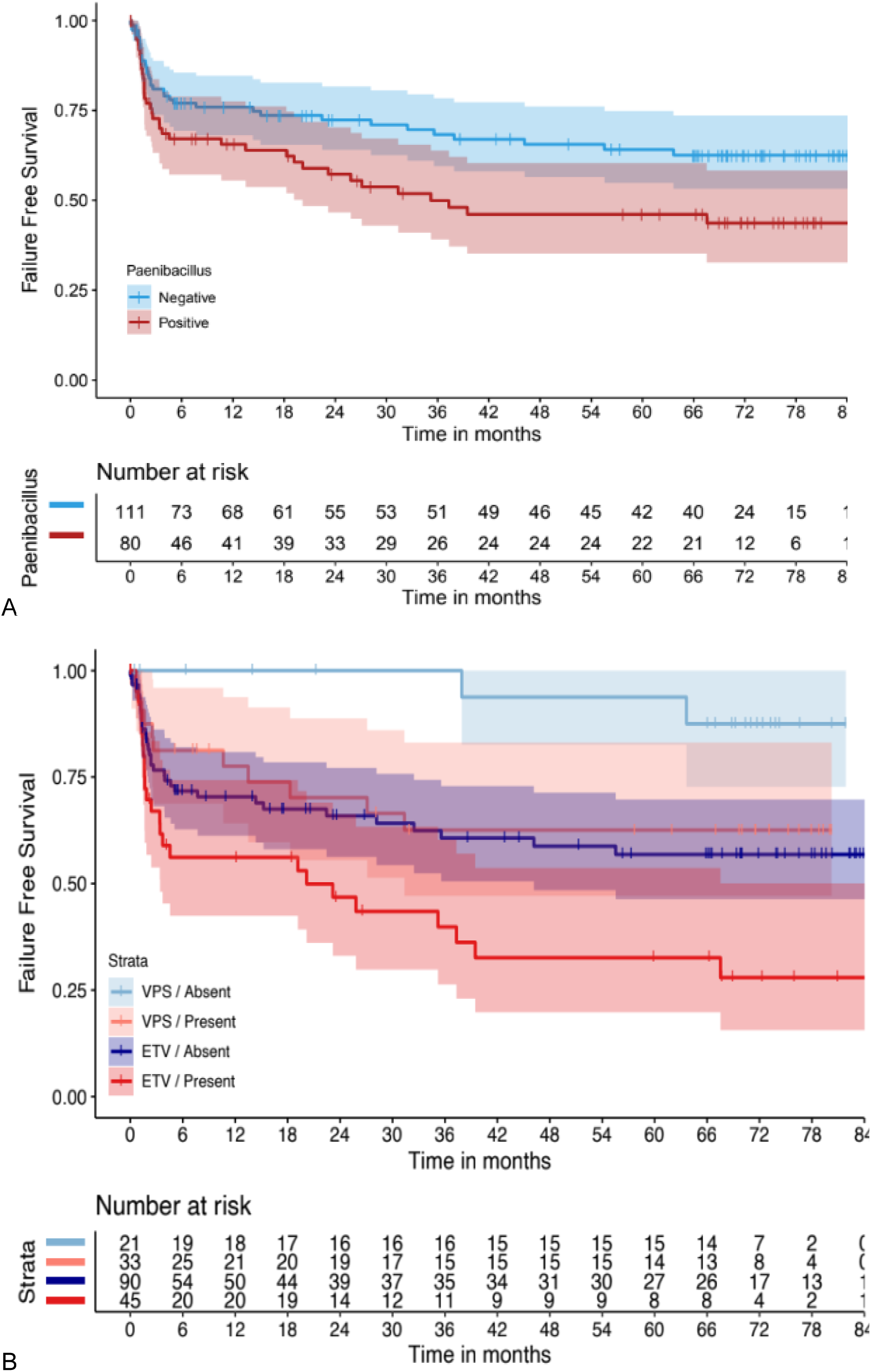
Time-to-event curves for diversion failure or death from any cause (A) overall and (B) stratified by diversion type.

**Figure 4.**
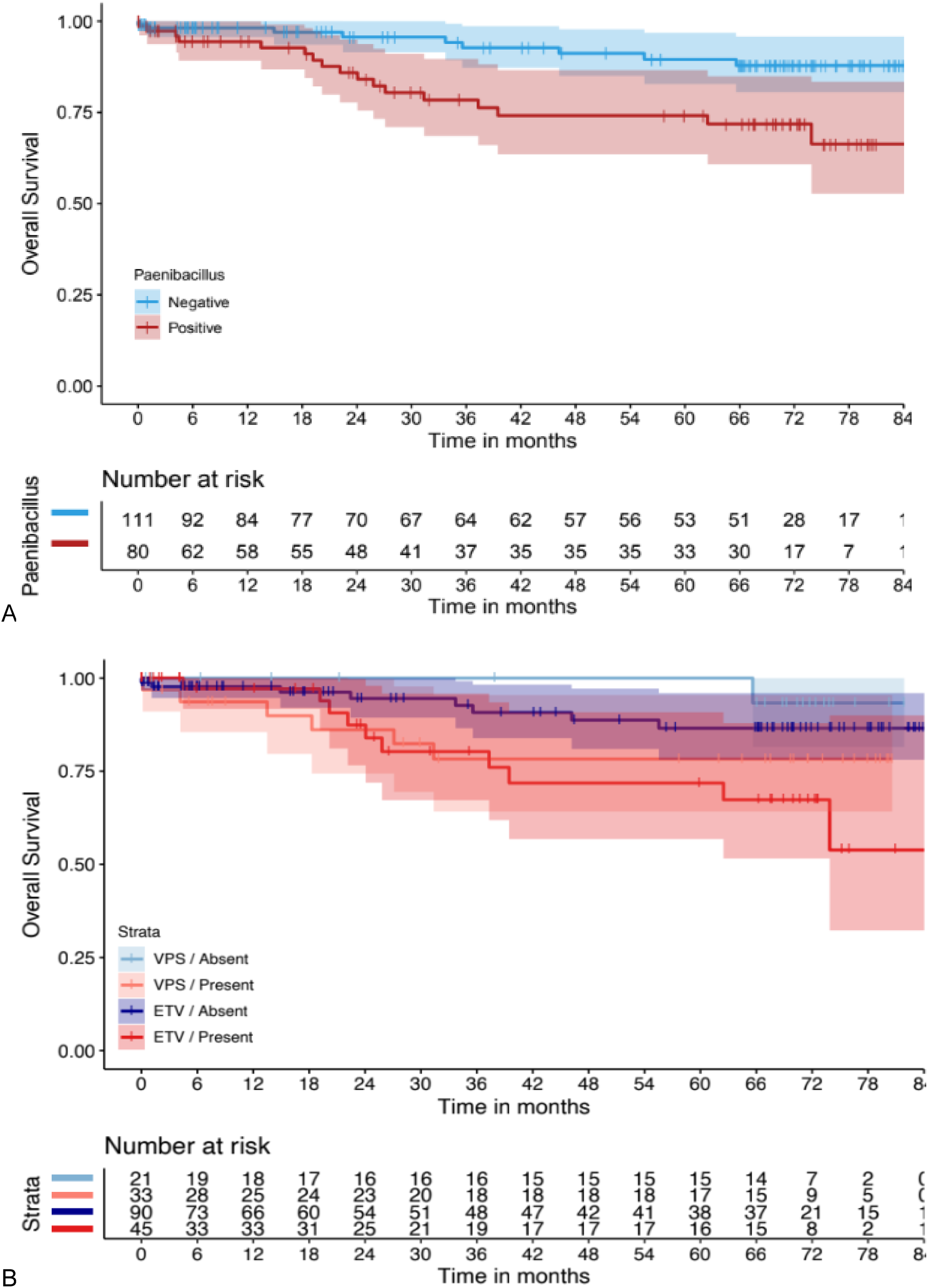
Time-to-event curves for death from any cause (A) overall and (B) stratified by diversion type.

**Figure 5.**
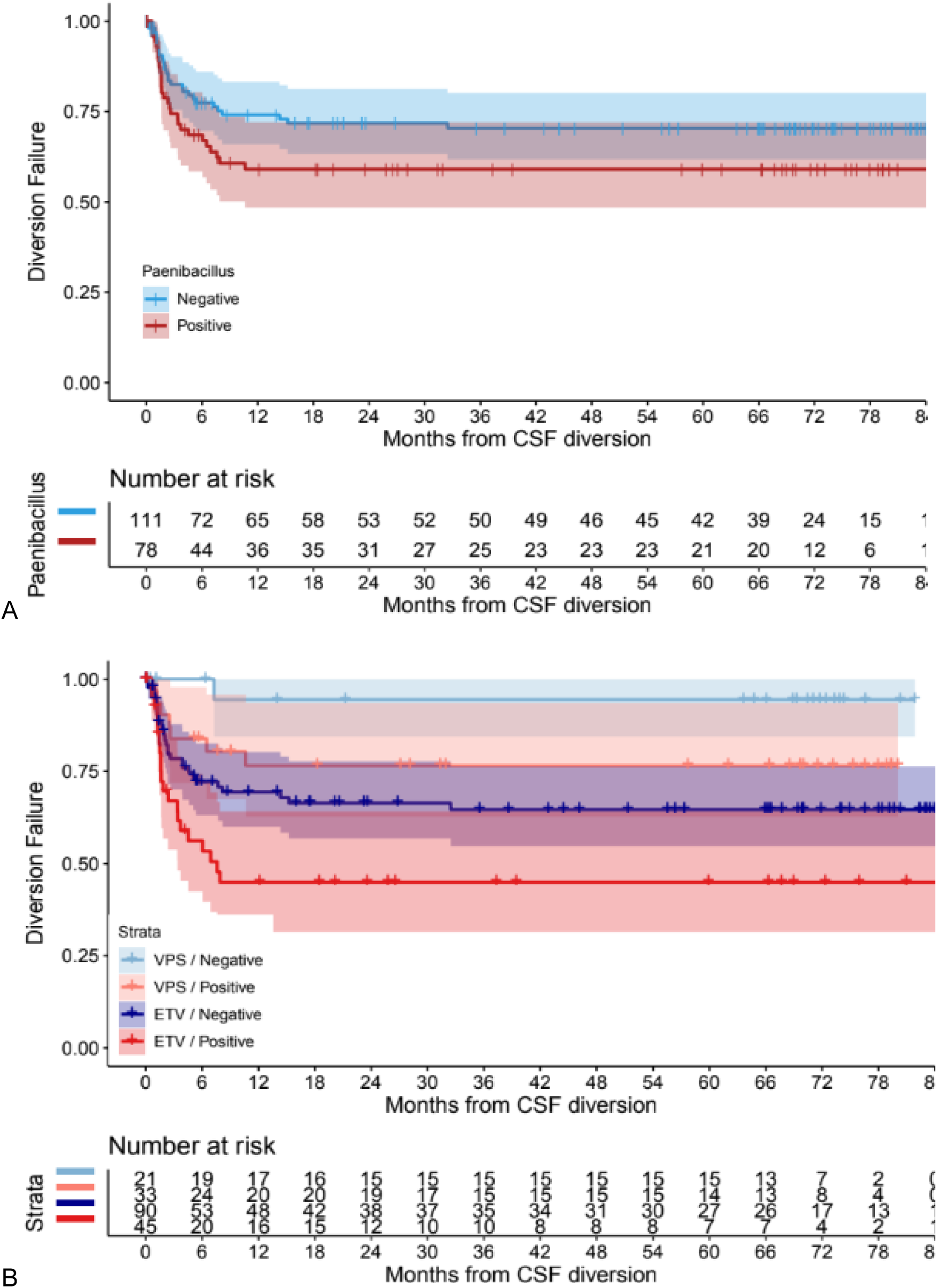
Time-to-event curves for diversion failure (A) overall and (B) stratified by diversion type.

**Table 2.** Comparison of outcomes at 6 and 12 months.

| Outcome | Comparison | 6-month RR (M-H) | 12-month RR (M-H) |
| --- | --- | --- | --- |
| **Composite Poor outcome** | VPS PP vs. PN | * | * |
|  | ETV PP vs. PN | 1.87 (0.91, 3.84) | 2.18 (1.15, 4.13) |
|  | PP ETV vs. VPS | 2.76 (1.07, 13.18) | 3.40 (1.26, 9.22) |
|  | PN ETV vs. VPS | * | * |
| **Mortality** | VPS PP vs. PN | * | * |
|  | ETV PP vs. PN | 2.15 (0.13, 34.4) | 2.11 (0.13, 33.8) |
|  | PP ETV vs. VPS | 0.40 (0.04, 4.4) | 0.40 (0.04, 4.39) |
|  | PN ETV vs. VPS | * | * |
| **Diversion failure** | VPS PP vs. PN | * | 5.23 (0.64, 42.5) |
|  | ETV PP vs. PN | 1.84 (0.98, 3.4) | 2.25 (1.26, 4.02) |
|  | PP ETV vs. VPS | 3.44 (1.27, 9.31) | 3.36 (1.43, 7.89) |
|  | PN ETV vs. VPS | * | 7.79 (1.06, 57.5) |

\*Not enough events to calculate.
Abbreviations: RR, relative risk; VPS, ventriculoperitoneal shunt; PP, Positive *Paenibacillus* polymerase chain reaction test result; PN, Negative *Paenibacillus* polymerase chain reaction test result; ETV, endoscopic third ventriculostomy

### Paenibacillus-Attributable Harm

After normalizing for diversion type, our cohort of 189 patients experienced 1.2 more diversion failures in the year after surgery and 7 more deaths than would be expected for a population this size without *Paenibacillus* infection. Ultimately, 8% of PIH patients experienced *Paenibacillus*-attributable harm.

### Prediction of ETV Success

As expected, in a post hoc analysis that included only patients who underwent an endoscopic diversion, patients with lower Uganda ETV success scores had higher rates of ETV failure (Table 3). No patients had a basic Uganda ETV success score >5; when an assessment of the aqueduct and the prepontine cistern were included, there were patients with scores across the whole range of possible scores. We found that *Paenibacillus* improved the ability of the basic Uganda ETV success score to predict ETV FFS and was also a useful addition to the extended Uganda ETV success score for the prediction of both FFS and diversion success. For example, an extended ETV success score in the middle tertile was associated with FFS, HR=1.64 (0.85, 3.21), but when the *Paenibacillus* PCR result was added to the model, a PP patient with an ETV success score in the middle tertile had a similar rate of FFS as a PN patient with a score in the lowest tertile, aHR=3.79 (1.58, 9.10) vs. aHR=4.01 (1.94, 8.31) (Supplemental Figure 2).

**Table 3.** Association of basic and extended Uganda ETV success scores with diversion failure-free survival with and without inclusion of *Paenibacillus* polymerase chain reaction result among patients who underwent ETV or ETV/CPC.

| Score | ETV success score |  | ETV success score + *Paenibacillus* PCR result |  |  | Likelihood ratio test |
| --- | --- | --- | --- | --- | --- | --- |
|  | Diversion failure or death<br>n/N (%) | aHR | Diversion failure or death<br>*Paenibacillus* negative<br>n/N (%) | Diversion failure or death<br>*Paenibacillus* positive<br>n/N (%) | aHR |  |
| Basic ETV success score* |  |  |  |  |  | 0.023 |
| 0-3 | 40/81 (49) | Reference | 20/49 (40) | 20/32 (62) | Reference |  |
| 4-6 | 18/53 (34) | 0.51 (0.27, 0.96) | 13/40 (32) | 5/13 (38) | 0.55 (0.29, 1.02) |  |
| 7-9 | - | - | - | - | - |  |
| Extended ETV success score† |  |  |  |  |  | 0.005 |
| 1-3 | 16/23 (70) | Reference | 9/15 (60) | 7/8 (88) | Reference |  |
| 4-6 | 21/48 (44) | 0.47 (0.23, 0.93) | 10/30 (33) | 11/18 (61) | 0.40 (0.20, 0.81) |  |
| 7-10 | 21/63 (33) | 0.28 (0.14, 0.58) | 14/44 (32) | 7/19 (37) | 0.25 (0.12, 0.52) |  |

\*Based on three criteria: Age: 0 if <6 months old, 1 if 6 months to <1 year, ≥3 if 1 year; Etiology of hydrocephalus: 1 if post-infectious, 2 if myelomeningocele, 0 if other; Choroid plexus cauterization: 0 if none, 2 if partial/unilateral, 4 if complete/bilateral
†Extended scoring system adds the status of the aqueduct at surgery: 0 if open, 2 if closed; and the extent of scarring in the prepontine cistern at surgery: 0 if scarred, 3 if unscarred.

### Sensitivity Analyses

Patients who underwent an ETV as their first surgical diversion procedure and were PP had a similar incidence of death or diversion failure as those who underwent ETV/CPC, 7/9 (78%) vs. 18/36 (50%), P=0.130, respectively. Similarly, overall survival was similar between PP patients who underwent ETV vs. ETV/CPC, 2/9 (22%) and 8/36 (22%), P=0.655, respectively. When adjusted for age at the time of surgery, a positive *Paenibacillus* CSF PCR was associated with FFS when either ETV, aHR=3.18 (1.03, 9.80), or ETV/CPC, aHR=2.41 (1.22, 4.75) was the type of endoscopic diversion performed. (Supplemental Figure 3).

Although no blood samples showed convincing transcripts of *Paenibacillus*, 13 CSF samples previously identified as positive using PCR demonstrated high levels of *Paenibacillus* transcripts (median = 4,564 reads, mean = 236,875.9 reads), indicating active infection in these patients. (Supplemental Figure 4).

## Discussion

Neonatal infections are the most common cause of infant hydrocephalus in many low-resource settings including Uganda.^1^ We describe a cohort of infants with PIH who experienced outcomes following surgical diversion similar to those previously reported.^2^ In this report, we sought to determine the impact of *Paenibacillus* detection in the CSF on surgical outcomes for postinfectious hydrocephalus. We found that *Paenibacillus* species detection in the CSF at the time of hydrocephalus surgery was associated with worse survival and neurosurgical outcomes than those without *Paenibacillus* detected and that this was true follow either VPS or ETV/CPC. These increased risks were greatest for infants who underwent an endoscopic diversion as their primary diversion surgery.

Infants who develop hydrocephalus following an early-life infection are typically thought to have PIH. Our findings suggest that some infants may have ongoing subacute or chronic active infection at the time they present with hydrocephalus. The possibility of active infection was confirmed by growth on culture in 3 cases and by detection of RNA transcript in 13 cases.^14^ Identification of a subacute infection at the time of neurosurgery allows the opportunity to provide antibiotic therapy which may improve the success of surgical diversion. This has previously been shown with hydrocephalus related to tuberculous meningitis; ETV outcomes are significantly improved if targeted antimicrobial therapy is initiated first.^20^ One possible explanation for the worse outcomes in the setting of *Paenibacillus* infection is that the ETV allows infected CSF to circulate through the brain, perhaps spreading the infection from one localized area to involve the whole brain.^21^ In contrast, VPS delivers infected CSF to the peritoneal cavity where *Paenibacillus* seems less likely to replicate efficiently. We have previously shown that peritoneal injections of *Paenibacillus* in a mouse model fail to establish active peritoneal infection.^14,22^ It is possible that subacute or chronic active infections due to other organisms may similarly affect the surgical outcomes of infants with presumed PIH. A thorough evaluation of the CSF for signs of infection including the use of molecular diagnostic studies when available may improve our understanding of why some children experience early diversion failure and allow opportunities to improve outcomes through antibiotic therapy and other interventions to treat the infection while also addressing the hydrocephalus.

ETV with or without CPC has become the preferred neurosurgical procedure to treat hydrocephalus in low-resource settings for several reasons: there is no implanted hardware that may malfunction,^23^ serve as a nidus for infection^24,25^ or cause other complications^26,27^ and there is a reduced risk of postoperative complications including over-drainage, intracranial hypotension and related subdural hemorrhage.^6,28^ Not all children with PIH are candidates for ETV since the anatomy must be conducive to being navigated by an endoscope and the floor of the third ventricle must be well-visualized.^29^ Our study was not adequately powered to detect differences in outcomes for ETV vs. ETV/CPC. However, our finding that *Paenibacillus* detection was associated with worse surgical outcomes was robust across the different surgical techniques. We identified a subset of patients who are at excess risk of ETV failure due to subacute infection at the time of CSF diversion. In fact, such PCR positive children should be considered to have ‘infectious hydrocephalus’ rather than PIH. It is therefore possible that identification and treatment of *Paenibacillus* infection at the time that an ETV is performed could represent a novel means of improving outcomes for infants with infectious hydrocephalus.

ETV success scores have been used in an attempt to improve the selection of patients for primary endoscopic diversion. Slight variations on the score have been used in different settings^30^; the Uganda ETV success score validated in sub-Saharan African infants and children includes the etiology of hydrocephalus (postinfectious vs. non-postinfectious), age, and the extent of choroid plexus cauterization performed.^7^ Adding a *Paenibacillus* PCR result to this score did not improve the ability of the score to accurately predict diversion failure; this was likely due to the small sample size and the homogeneity within the cohort with respect to the scores assigned for age (all of our patients were <6 months of age) and hydrocephalus etiology. *Paenibacillus* status may be found to be an impactful addition to the basic ETV success score once it is evaluated in a larger, more diverse, cohort of patients. However, we did find that adding the *Paenibacillus* PCR result to the model increased the ability of the extended Uganda ETV success score to predict both diversion success and FFS; a positive PCR result functionally changed the score to the next lower tertile. For example, a patient with a Uganda ETV success score of 4-6 who also had a positive CSF *Paenibacillus* PCR result had the same rate of FFS as a patient with a Uganda ETV success score of 1-3. Importantly, unlike factors such as the extent of CPC, or, in the absence of preoperative MRI, state of the aqueduct or extent of cisternal scarring, the *Paenibacillus* PCR result can be determined preoperatively if a ventricular puncture is performed to sample the CSF. Adding a *Paenibacillus* CSF PCR to the preoperative laboratory assessments of infants with hydrocephalus could allow for more refined prediction of the likelihood of treatment success. Perhaps more importantly, this might offer the opportunity to improve the outcome with concurrent antibiotic treatment. It is conceivable that the likelihood of ETV/CPC success could be increased through antibiotic therapy initiated prior to or at the time of the surgical diversion.

Optimal antibiotic treatment and the timing of primary CSF diversion in the setting of *Paenibacillus* CNS infection is not known but antibiotic susceptibility testing results of available isolates suggests that vancomycin and other commonly used antibiotics may be ineffective.^22,31–33^ Our results suggest the possibility that the right antibiotic treatment protocol could positively affect both treatment outcome for ETV/CPC and VPS and overall survival among infants with PP PIH. Based on our findings, we estimate that 8% of Ugandan infants with PIH experienced “*Paenibacillus*-attributable harm” that could potentially be prevented with effective antibiotic treatment. When we consider that PIH is a common cause of morbidity and mortality in this part of the world, the clinical implications of this finding are quite large and could impact hundreds of children each year. Future clinical trials should include a comparison of surgical outcomes following antibiotic treatment.

### Limitations

Infants included in this analysis were considered to have PIH. However, PIH was defined based on clinical history and imaging findings consistent with prior infection without evidence of congenital malformations. Some infants may have signs of infection without actually having infection of the CNS and could have been misclassified as having PIH. The majority of infants included in the current study were not treated with a course of antibiotics at the time of their diversion surgery. The PCR results were performed on stored samples and the results were not available in real time.

## Conclusion

PP infants with hydrocephalus are at increased risk of death and diversion failure. Pre-operative testing for *Paenibacillus* would allow for antibiotic therapy to be provided which may significantly improve outcomes. Extra care should be taken in choosing a surgical technique for infants with PP infectious hydrocephalus.

## Data Availability

All data produced in the present study are available upon reasonable request to the authors

---

**Supplemental Table 1.** Sensitivity, specificity, positive predictive value and negative predictive value of computed tomography scan findings for diagnosing *Paenibacillus* infection in infants with hydrocephalus.

| Number of CT scan findings* | Sensitivity | Specificity | Positive Predictive value | Negative predictive value |
| --- | --- | --- | --- | --- |
| 0 | 5.1% (2.0, 8.3) | 79.3 (73.5, 85.1) | 14.8 (9.8, 19.9) | 54.3 (47.2, 61.4) |
| 1 | 15.4 (10.2, 20.5) | 70.3 (63.8, 76.8) | 26.7 (20.4, 33.0) | 54.2 (47.0, 61.3) |
| 2 | 20.5 (14.8, 26.3) | 80.2 (74.5, 85.9) | 42.1 (35.1, 49.1) | 58.9 (51.9, 66.0) |
| 3 or 4 | 59.0 (52.0, 66.0) | 70.3 (63.8, 76.8) | 58.2 (51.2, 65.3) | 70.9 (64.4, 77.4) |

\*Presence of abscess, calcifications, loculations or intraventricular debris

**Supplemental Figure 1.**
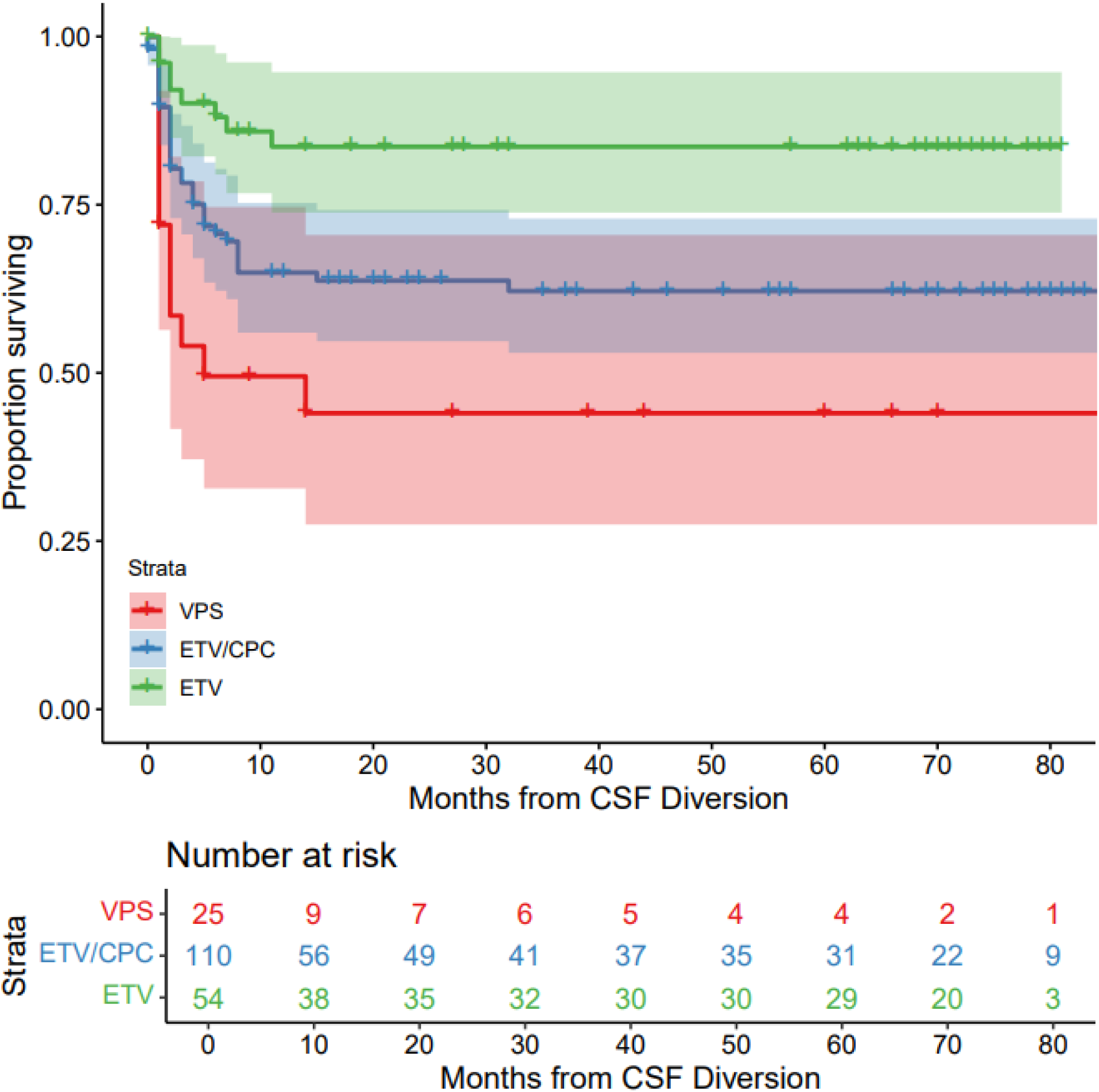
Diversion failure-free survival by surgery type.

**Supplemental Figure 2.**
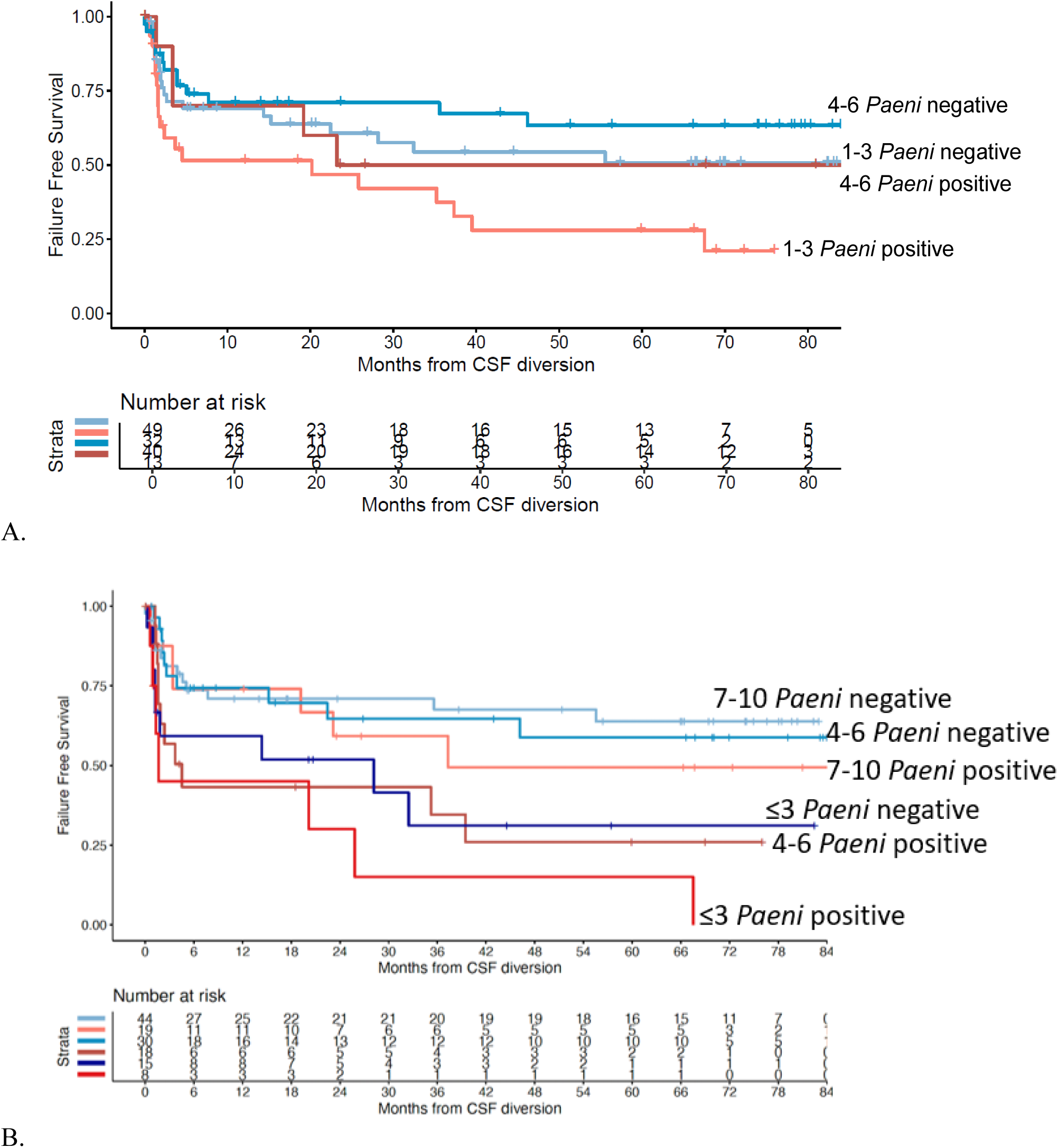
Time to diversion failure or death from any cause for patients who underwent endoscopic diversion stratified by *Paenibacillus* PCR result and the Uganda ETV success score (A) or extended Uganda ETV success score (B).

**Supplemental Figure 3.**
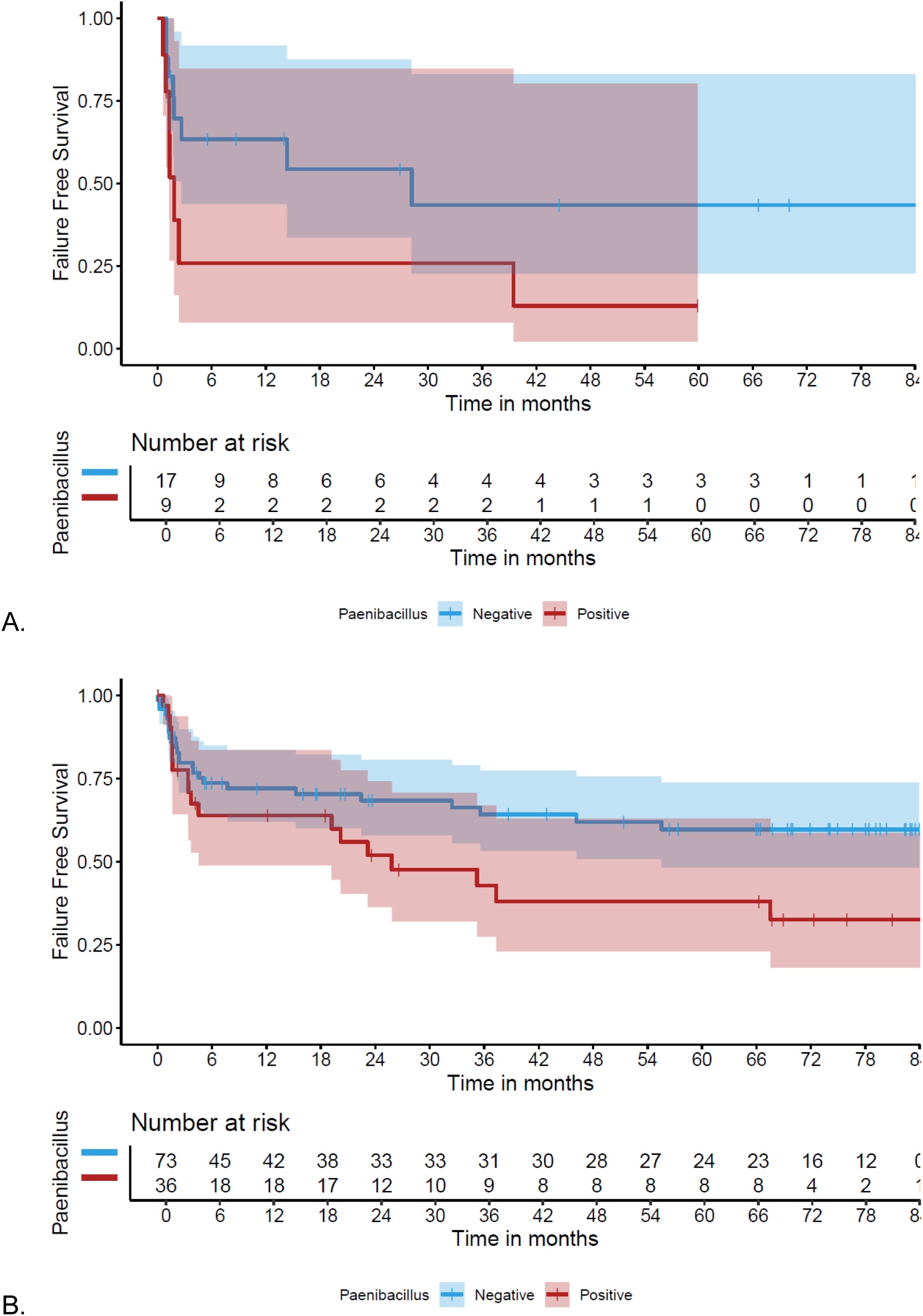
Time-to-event curves for death or diversion failure for infants who underwent an endoscopic third ventriculostomy (A) or endoscopic third ventriculostomy with choroid plexus cauterization (B).

**Supplemental Figure 4.**
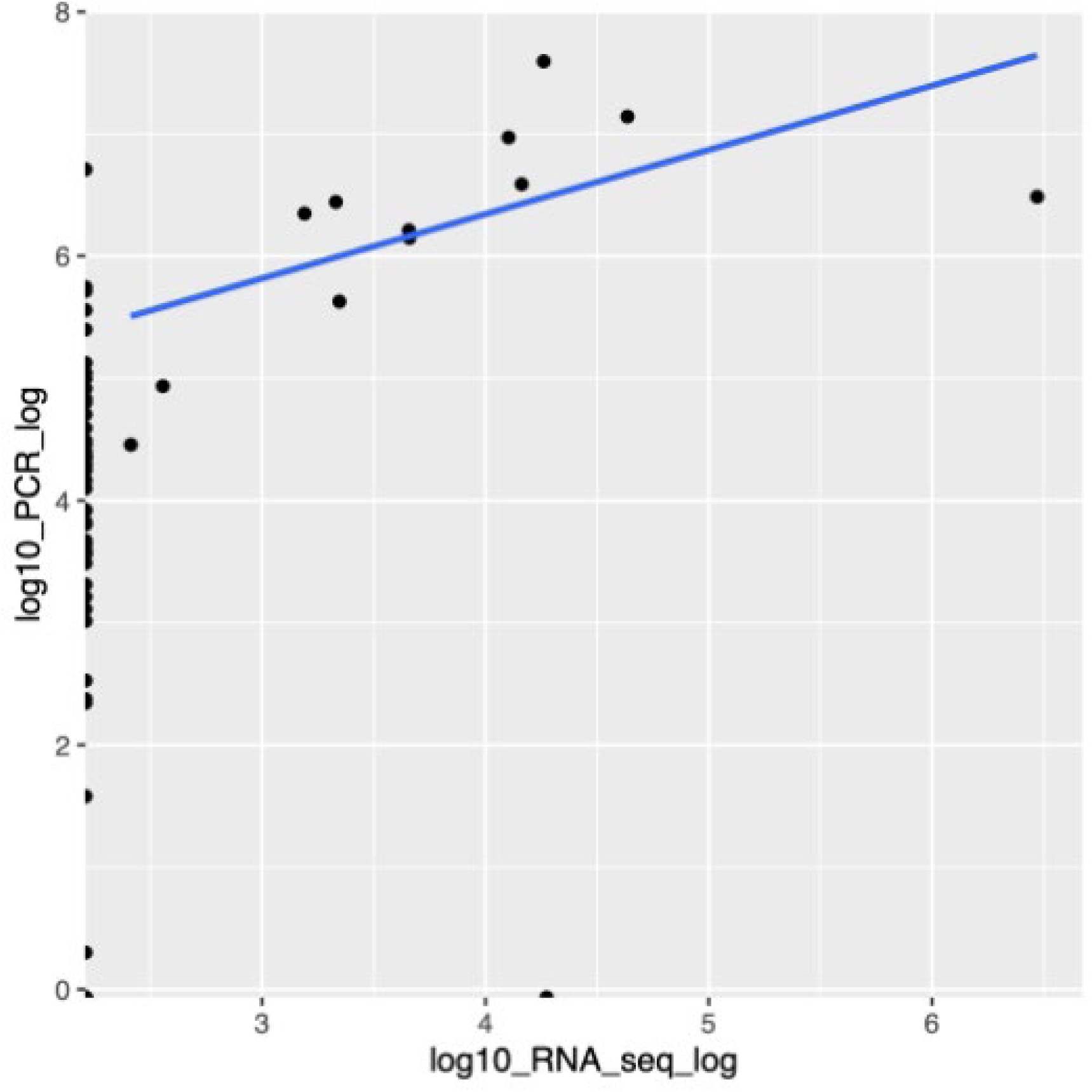
Concentrations of *Paenibacillus* messenger RNA transcript detected in the cerebrospinal fluid of patients with a cerebrospinal fluid quantitative PCR positive for *Paenibacillus*.

## Notes

Conflicts of interest: All authors have no conflicts to report.

### Competing Interest Statement

The authors have declared no competing interest.

### Funding Statement

This study and the prior prospective study were funded by the National Institutes of Health under NIH Directors Pioneer Award 1DP1HD086071 (S.J.S), NIH Directors Transformative Award 1R01AI145057 (S.J.S), as well as NICHD 5R01HD085853 (B.C. W., A.V.K., S.J.S).

### Author Declarations

The study protocol was approved by the Institutional Review Boards at CURE Childrens Hospital of Uganda with oversight of the Ugandan National Council on Science and Technology, The Pennsylvania State University, and Yale University.

